# Impact of Vitamin D supplementation on cognition in adults with mild to moderate vitamin D deficiency: Outcomes from the VitaMIND randomised controlled trial

**DOI:** 10.1101/2024.11.21.24317708

**Authors:** Anne Corbett, Rod Taylor, David Llewellyn, Janice M. Ranson, Adam Hampshire, Ellie Pickering, Abbie Palmer, Dag Aarsland, Dorina Cader, Diana Frost, Clive Ballard

## Abstract

**Background:** Preserved cognitive health with ageing is a public health imperative. Vitamin D deficiency is associated with poor cognition, but it is unclear whether supplementation would provide benefit, particularly in individuals with mild/moderate deficiencies which do not have other clinical risks. The objective of this study was to establish the impact of daily vitamin D supplementation on cognition in older adults with mild to moderate vitamin D deficiency.

**Methods and Findings:** Two-arm parallel 24-month randomised controlled trial, with Vitamin D supplementation compared with a placebo. This was a remote trial, completed from home involving 620 adults 50 years or older with mild to moderate vitamin D deficiency and early cognitive impairment. The primary outcome was executive function measured through Trail making B and other secondary measures of cognition, function and wellbeing.

Vitamin D supplementation conferred no significant benefit to executive function compared to placebo at follow-up on the primary outcome (between-group difference: 5770, 95% CI: -2189 to 13730) or cognition, function, or wellbeing. Secondary analyses in defined subgroups and a per-protocol analysis also showed no significant impact on any outcome measures.

**Conclusions:** Vitamin D supplementation produced no measurable improvement in cognitive outcomes in older adults with mild to moderate vitamin D deficiency. The remote trial methodology provides an innovative approach to large-scale trials.

**Trial Registration:** ISRCTN79265514 https://www.isrctn.com/ISRCTN79265514

## Introduction

Dementia is a devastating condition that affects over 850,000 people in the UK and carries enormous emotional, societal and financial burdens, with an estimated cost of £26.3 billion each year to the UK health service(1). Worldwide, this figure reaches US$818billion. While new disease-modifying treatments are in the pipeline, there is a continuing imperative to deliver approaches to reduce the risk of cognitive decline and dementia, particularly in high-risk groups. The largest at-risk group for dementia are individuals with Mild Cognitive Impairment (MCI), which affects 10-20% of older adults, of whom up to 10% develop dementia each year(2). A larger and less well-defined group of individuals experience Age-Associated Cognitive Decline (AACD), a risk state for MCI, which has been clinically defined by recent diagnostic criteria(3). The opportunity to apply precision medicine approaches to risk reduction in these groups would enable tailored treatment and preventative interventions to be targeted in the most impactful way to achieve better public health. The potential impact of such a strategy to preserve cognition in older age and delay the onset of dementia symptoms, even by a few months, could have a significant impact at a population level and improve the health and wellbeing of older adults.

There is good evidence to support several approaches for reducing the risk of dementia. One promising avenue is through supplementation of dietary deficiencies, particularly with Vitamin D. Vitamin D deficiency is prevalent in the UK, particularly in the winter months. The Scientific Advisory Committee on Nutrition reports that 23% of adults fulfil criteria for severe vitamin D deficiency, categorised as clinically at risk of reduced bone mineralisation, causing bone pain, muscle pain and fatigue, and potential impacts on respiratory and cardiovascular conditions (< 25nmol/L of 25-hydroxy-vitamin D (25(OH)D) in the blood), while 40.4% fulfil criteria for inadequate levels (<50nmol/l)(4-6). These figures highlight the prevalence of deficiency across the UK population. Seasonality is a significant factor in levels, with severe deficiency prevalence rising from 23% to 29% in adults over 65 from summer to winter months. Dietary supplementation, therefore, offers a means of significantly increasing vitamin D levels above the current UK baseline.

A rapidly growing body of evidence indicates that Vitamin D may play a role in brain health and cognition. Vitamin D receptors (VDR), one of the nuclear receptor family of transcription factors, are found extensively throughout the brain, and they are associated with neuroprotection and anti-inflammatory effects(7). For example, vitamin D reduces the hallmarks of Alzheimer’s disease, including amyloid beta in in vitro models(8, 9). Vitamin D was first identified as a potential intervention for dementia prevention in 2009 by a study that linked low vitamin D levels in 1,766 older adults to cognitive impairment(10). Subsequently, a large prospective study established that severe vitamin D deficiency (<25 nmol/L) was associated with a risk of substantial cognitive decline over six years by around 60%, and a more recent study showed that vitamin D exposure was associated with a significant reduction in dementia risk, particularly in individuals at greater risk of dementia(11). People with severe vitamin D deficiency are 120% more likely to develop dementia over six-years(12). These landmark studies have been replicated by other groups and confirmed in a series of systematic reviews and meta-analyses(13, 14). This evidence base, in addition to clinical links to conditions such as osteoporosis and osteoarthritis, clearly shows the importance of treating severe vitamin D deficiency to improve cognitive health. The strength of this evidence would preclude a randomised trial in these patients since the benefit to health is so clear. However, the case is less clear-cut with regard to mild and moderate deficiency, where a trial is warranted. A meta-analysis of 37 studies explored cognitive performance in people above and below the mild deficiency threshold of 50nmol/l and reported significantly worse cognition in individuals with mild vitamin D deficiency. A further observational and Mendelian randomisation study also showed non-linear associations of dementia risk in individuals up to 50nmol/l(15, 16) Therefore, the key emerging question is whether treating mild to moderate vitamin D deficiency confers benefits regarding cognitive outcomes.

People with severe vitamin D deficiency are usually identified and treated through established clinical care pathways, ensuring that their resulting dementia risk and other health issues are likely to be mitigated through effective supplementation. However, mild to moderate deficiencies of 25-50nmol/l are less likely to be detected despite being prevalent in the community. There is a need to establish whether supplementation of these groups could improve cognitive outcomes and whether this should be incorporated into risk reduction strategies for wider public health initiatives. This study conducts a definitive trial to determine whether Vitamin D supplementation is an effective means of maintaining cognitive function in people with mild to moderate vitamin D deficiency. Trials of this nature, which are investigating low-risk interventions in a large cohort of individuals in the community, offer the opportunity to employ innovative remote trial methodologies, using a combination of online assessment, digital tools for engagement, and remote dispensing approaches to deliver large-scale trials without the need for in-person clinic visits and assessments. The PROTECT-UK platform offers the infrastructure to deliver remote, virtual trials(17). This study represents the first large-scale proof of concept for this model of trial delivery.

## Methods

### Study Design

This VitaMIND study was a parallel, double-blind, 24-month remote randomised controlled trial to establish the impact of vitamin D supplementation on cognition, function and wellbeing in adults over 50 with mild to moderate vitamin D deficiency and age-associated cognitive decline. The study was approved by the Wales Research Ethics Committee 3 under the UK Health Research Authority (Ref 19/WA/0007). The protocol is registered on the ISRCTN database (Ref: ISRCTN79265514). The study is reported in accordance with CONSORT guidelines (ref). Patients, clinicians, and the research team were all blinded to group allocation.

### Participants

Adults over the age of 50 in the UK were invited to take part through the UK online ageing cohort, PROTECT(17, 18). All participants were already registered on the PROTECT cohort and were invited to the trial by email correspondence as part of the consent for contact in place in the PROTECT cohort. Eligible participants were over the age of 50, without a diagnosis of dementia, had access to a computer and the internet, fulfilled criteria for high risk of vitamin D deficiency of 50nmol/l 25(OH)D or below based on a self-reported screening questionnaire (Table1) and fulfilled criteria for AACD (performing one standard deviation below norm on at least one cognitive test in the PROTECT-UK Cognitive Test System, described elsewhere)(19). The PROTECT-UK cohort was pre-screened for age and AACD criteria. Interested individuals registered and provided consent for the study through an ethically approved digital consent process embedded on the PROTECT website. Participants then accessed the trial by navigating to the VitaMIND trial area on their online dashboard. Automated emails were scheduled to remind participants to take their study tablets and complete their cognitive assessments.

### Screening Questionnaire for Vitamin D deficiency

In order to identify participants with Vitamin D deficiency (<50 nmol/L), predictive algorithms were trained using data from 3,519 community-dwelling participants aged 50 and over from the English Longitudinal Study of Aging (ELSA)(20). Potential predictors were based on those identified in previous risk prediction models for vitamin D deficiency, as well as those with an established association with vitamin D levels. In the absence of a suitable external test dataset, split-sample development and validation were conducted (80% used for development, 20% for validation). Missing data was dealt with using multivariate imputation by chained equations. Predictors were selected using multivariable fractional polynomial bootstrapped regression with 1000 bootstrap replications. Ten predictors were included in the final algorithm: Body mass index, treatment for osteoporosis, month of assessment, smoking status, exercise, marital status, age, alcohol consumption, ethnicity and self-rated poor health. See Table 1 for full details of the questionnaire and response options. Participants with a 30% or higher predicted probability of mild Vitamin D deficiency (<50 nmol/L) were eligible for the study.

**Table 1:** Screening questionnaire and algorithm for vitamin D deficiency.

| Description | Wording | Response options | Notes |
| --- | --- | --- | --- |
| **BMI** | How much do you weigh?<br>How tall are you? | Kg or stones and pounds<br>Meters or feet and inches | **To calculate BMI**<br>**Either:**<br>Weight (kg) / height (m)<sup>2</sup><br>**Or:**<br>Weight (lb) / height (in)*703<br>(in which case, weight first needs to be converted to lbs and height converted to inches. |
| **Treatment for osteoporosis** | Do you take calcium pills or Vitamin D for osteoporosis, sometimes called thin or brittle bones? | Yes / No | 'No' is the reference category |
| **Month of assessment** | N/A | N/A |  |
| **Physical activity** | A. Do you take part in sports or activities that are vigorous?<br>B. Do you take part in sports or activities that are moderately energetic?<br>C. Do you take part in sports or activities that are mildly energetic? | A1 More than once a week<br>A2 Once a week<br>A3 1-3 times a month<br>A4 Hardly ever or never<br>B1 More than once a week<br>B2 Once a week<br>B3 1-3 times a month<br>B4 Hardly ever or never<br>C1 More than once a week<br>C2 Once a week<br>C3 1-3 times a month<br>C4 Hardly ever or never | Physical activity has four categories depending on combination of responses to these three questions:<br>'0' is the reference category<br>A participant is assigned to the lowest number category for which they are eligible.<br>Category response requirements are below:<br>0 = High (A1)<br>1 = Moderate (B1 or B2) or (A2 or A3)<br>2 = Low (C1 or C2) or (B2 or B3 or B4) + (A4)<br>3 = Sedentary (C3 or C4) + (B4) + (A4) |
| **Current smoking status** | Do you currently smoke cigarettes, cigars or a pipe? | Yes / No | 'No' is the reference category |
| **Age** | What is your date of birth? | Date / Month / Year |  |
| **Couple status** | Are you currently married or living with a partner? | Yes / No | 'Yes' is the reference category |
| **Ethnicity** | To which of these groups | White / Mixed / | 0/1 binary variable |
|  | do you consider that you belong? | Black / Black British / Asian / Asian British / Other | 'White' versus all other categories. 'White' is the reference category. |
| **Weekly alcohol consumption** | During the last seven days, how many measures of spirits did you have? | Continuous | The responses to these three questions need to be totalled to produce a single weekly number of drinks. |
|  | During the last seven days, how many glasses of wine did you have? | Continuous |  |
|  | During the last seven days, how many pints of beer did you have? | Continuous |  |
| **Self-rated poor health** | Would you say your health is... | Excellent / Very good / Good / Fair / Poor | 0/1 binary variable 'Poor' versus all other categories. 'Other' is the reference category. |
| **Algorithm to predict Vitamin D deficiency (<50nmol/l)**<br>$\text{gen pred\_vitd50} = -2.987615 + (\text{bmi} \times .0642204) + (\text{osteomed} \times -1.178731) + (((\text{month}/10)^3 - .4303164208) \times -1.336699) + (((\text{month}/10)^3 \times \ln(\text{month}/10) + .1209525476) \times 7.583981) + (\text{smoker} \times .7837381) + (\text{exercise\_1} \times .4699397) + (\text{exercise\_2} \times .7609146) + (\text{exercise\_3} \times .7766721) + (((\text{age}/10)^2 - 44.36322732) \times -.1511723) + (((\text{age}/10)^3 - 295.4844962) \times .0147316) + (\text{couple} \times .4051173) + (\text{ethnic} \times 1.224978) + (((\text{drink}+1)/10)^{-.5} - 1.226242542) \times .1254607) + (\text{selfhealthpoor} \times .1064709)$<br>$\text{Gen prob\_vitd50} = (\exp(\text{pred\_vitd50}) / (1 + \exp(\text{pred\_vitd50}))) \times 100$ | | | |

### Treatment Interventions

Participants were randomised to receive either 4000IU Vitamin D3, supplied in two-piece HPMC capsules, or an identical placebo capsule without the active Vitamin D3 supplement. This dose is the highest publicly available vitamin D3 supplement and was chosen for relevance to existing available treatments and due to known safety data. Capsules were provided in blister packs of 28, stamped with the days of the week to aid compliance. Participants were instructed to take one tablet daily for the 24-month duration of the study.

### Outcome Measures

Outcome measures were completed at baseline, six, 12 and 24 months. All outcome measures were completed online through the PROTECT cohort platform.

### Primary Outcome Measure

The primary outcome measure was executive function and task-switching as measured by a computerised version of the well-validated Trail making B task in which a participant connects an alternating sequence of alphanumeric characters(21). Total time and accuracy are captured and combined to provide a total score.

### Secondary Outcome Measures

Secondary cognitive outcomes were measured using a wider computerised cognitive test system consisting of five cognitive tests described in full in previous papers (22) (18, 23). The test system included measures of spatial working memory (Paired Associate Learning, Self-Ordered Search), numerical working memory (Digit Span) and executive function (Verbal Reasoning, Switching Stroop). Additional secondary outcome measures were also collected to assess function (Instrumental Activities of Daily Living), behaviour (Mild Behaviour Impairment Scale) and wellbeing (EQ5D)(24-26).

### Sample Size

The sample size calculation was based on a published study of online cognitive training in adults over 50, which showed significant change in executive function in adults with AACD (27). Based on an effect size of 0.3, which is clinically meaningful and has been achieved through similar trials in this field, 165 participants would be required for each group to provide 80% power at a two-sided 0.05 significance level, assuming a conservative drop-out rate of 40%. Therefore, a total of at least 462 participants (231 per group) were required for this trial.

### Randomisation and masking

Randomisation of participants was achieved through a purpose-built algorithm embedded in REDCap Cloud. Randomisation occurred after a participant had completed their baseline assessments. The algorithm allocated participants randomly, stratifying for age (age brackets of five years), gender and Vitamin D deficiency severity (mild-moderate (< 50 nmol/L) or severe (< 25 nmol/L) Vitamin D level).

The treatment allocation of participants was recorded electronically in REDCap Cloud, hosted by the University of Exeter’s Clinical Trials Unit. This ensured that both participants and the research team were blind to allocation, thus removing any bias. Study staff with direct participant contact were not involved in any data analysis, thus avoiding any unconscious bias as a result of their contact.

### Safety Monitoring

All AEs and SAEs occurring from the time of the start of trial treatment until the end of the trial were reported by participants using an online form. Participants were prompted to enter any details of events at each quarterly login event and encouraged to report any event proactively using the online form or by contacting the study team by telephone and email. All AEs were logged and reported to the study team for review within 24 hours.

### Compliance

Participants completed a regular tablet count record on the study website to track compliance across the full study and to support the per-protocol analysis. Ten per cent of the cohort was also randomly selected to complete finger-prick blood tests at baseline and 24 months to quantify vitamin D blood levels. Participants received the vitamin D blood spot test kit in the post, completed the tests at home according to the kit instructions, and returned them to the Sandwell and West Birmingham NHS Foundation Trust laboratory for analysis.

### Data Analysis

Analysis was performed according to a prespecified statistical analysis plan. The primary analyses compared primary and secondary outcomes between intervention and control groups at 24 months using linear regression models with adjustment for baseline outcome score and stratification variables. Primary analyses were based on an intention-to-treat (ITT) approach.

A number of secondary analyses were undertaken. A repeated measures analysis using a mixed effects linear regression model with a random effect on participants) for primary and secondary continuous outcomes, to compare intervention and control groups including data from participants with observed data for at least one of the three follow-up timepoints. A fixed effect interaction between the time point and trial arm was used to evaluate differential treatment effects across time points. Adjustments for baseline covariates were made for the primary analysis regression models. Multiple imputation using chained equations was used to impute missing primary and secondary continuous outcome data. Imputation models were informed by the treatment arm, baseline scores, and stratification covariates to be included in the primary analysis model. Primary analysis models for primary and secondary outcomes were rerun using imputed data sets. To assess the potential effect of tablet adherence, an additional analysis was undertaken with the primary ITT analysis adjusted for the difference in baseline and 24-month follow-up tablet count. Blood test data for compliance was analysed using a t-test.

## Results

### Cohort Characteristics

620 participants consented to the study between 1^st^ September 2020 and 26 ^th^ March 2021, of whom 75% were female, with an average age of 60 (SD 6.58) and an average educational attainment corresponding to completion of secondary education with an additional vocational qualification. 310 participants were randomised to the Vitamin D intervention group, and 310 were randomised to the control group. There were no significant differences between the characteristics of the two groups. In the sub-group of participants who completed a blood sample at baseline, the median vitamin D level was 42 nmol/l with no significant difference between groups (p=0.44). The baseline characteristics of the study participants are described in Table 2, and the flow of participant selection through the study is presented in Figure 1. The trial ended after the last follow-up assessment of the last participant was completed.

**Table 2:** Cohort characteristics for the VitaMIND trial.

| Characteristic |  | Full cohort<br>n = 620 | VitaMIND group<br>n = 310 | Control group<br>n = 310 |
| --- | --- | --- | --- | --- |
| **Age** | Range | 50 – 80 | 50 – 80 | 50 – 80 |
|  | Mean (SD) | 60.7 (6.58) | 60.3 (6.27) | 60.8 (6.73) |
| **Sex** | Male n (%) | 154 (25%) | 75 (25%) | 79 (25%) |
|  | Female n (%) | 466 (75%) | 235 (75%) | 231 (75%) |
| **Educational attainment** | Mean (SD) | 3.17 (1.38) | 3.19 (1.35) | 3.14 (1.41) |
| **Ethnicity** | White n (%) | 608 (98) | 301 (97) | 303 (98) |
|  | Mixed n (%) | 4 (<1) | 3 (<1) | 1 (<1) |
|  | Asian n (%) | 7 (<1) | 3 (<1) | 4 (<1) |
|  | Black n (%) | 1 (<1) | 1 (<0.5) | 0 (0) |

**Fig 1.**
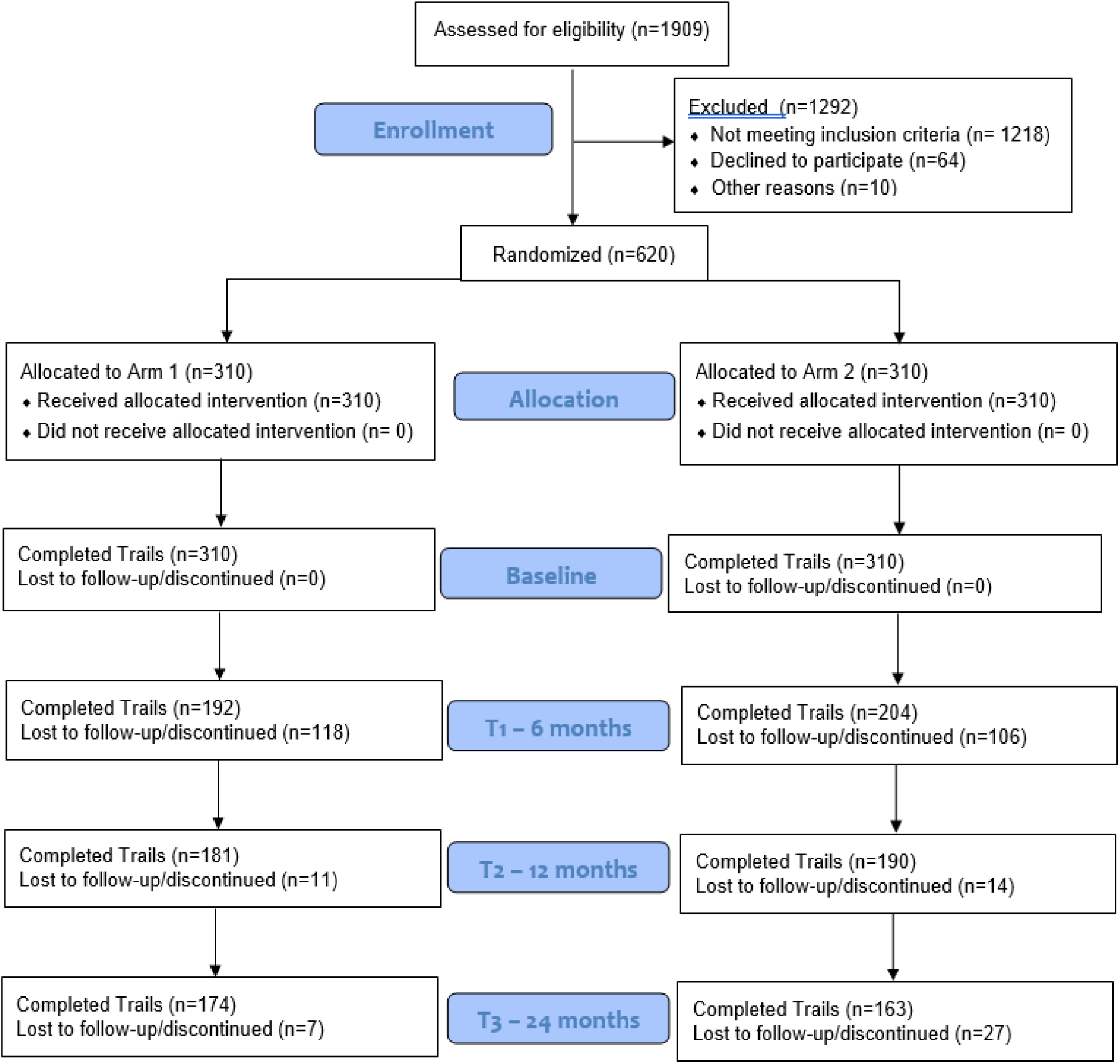
CONSORT chart showing the flow of participants through the VitaMIND trial.

### Impact of Vitamin D on cognitive outcomes

Analysis showed no significant benefits to cognition in the Vitamin D intervention group compared to placebo at 24 months. In the primary outcome of executive function measured by the Trail making task, the ReaCT intervention conferred no significant benefit compared to the control task (Cohen’s D Effect Size (ES) 0.11; P=0.16) (Table 3). In the secondary outcome measures, the treatment group also showed no significant benefit compared to the control group in numerical working memory (ES 0.08; P=0.38)), spatial working memory (Paired Associate Learning P=0.84; Self-Ordered Search P=0.64) or executive function (Switching Stroop P=0.86; Verbal Reasoning P=0.92) (Table 3).

**Table 3:** Impact of Vitamin D supplementation on executive function (primary outcome), cognition (Secondary outcomes), function, behaviour and wellbeing at 24 months.

|  | Vitamin D Group |  | Control Group |  | Between group difference* |  | Effect size |
| --- | --- | --- | --- | --- | --- | --- | --- |
|  | Baseline | 2-years | Baseline | 2-years |  |  |  |
|  | N mean (SD) | N mean (SD) | N mean SD | N mean SD | Mean (95% CI), | p-value |  |
| Primary outcome |  |  |  |  |  |  |  |
| Trail making B | 310<br>61869<br>(27299) | 174<br>61388<br>(22976) | 310<br>61421<br>(31949) | 163<br>55954<br>(49506) | 5770 (-<br>2189 to<br>13730) | 0.16 | 0.116552 |
| Secondary cognitive outcomes |  |  |  |  |  |  |  |
| Baddeley Grammatical Reasoning | 310<br>36.0<br>(11.7) | 169<br>37.8<br>(11.4) | 310<br>36.7<br>(11.8) | 150<br>37.7<br>(10.8) | 0.1<br>(-1.4 to<br>1.5) | 0.92 | 0.009259 |
| Switching Stroop | 310<br>40.2<br>(17.6) | 174<br>44.5<br>(17.2) | 310<br>41.5<br>(17.0) | 163<br>43.2<br>(18.6) | 0.7<br>(-2.2 to<br>3.7) | 0.63 | 0.037634 |
| Paired Associate Learning | 310<br>3.98<br>(1.02) | 170<br>4.11<br>(0.92) | 310<br>4.04<br>(1.18) | 155<br>4.17<br>(0.95) | -0.02<br>(-0.21 to<br>0.17) | 0.84 | 0.021053 |
| Digit Span | 310<br>7.32<br>(1.64) | 166<br>7.51<br>(1.55) | 310<br>7.23<br>(2.00) | 149<br>7.39<br>(1.39) | 0.11<br>(-0.14 to<br>0.38) | 0.38 | 0.079137 |
| Self-Ordered Search Task | 310<br>7.54<br>(1.98) | 172<br>7.62<br>(1.80) | 310<br>7.59<br>(2.13) | 157<br>7.59<br>(1.91) | 0.09<br>(-0.28 to<br>0.45) | 0.64 | 0.04712 |
| Secondary non-cognitive outcomes |  |  |  |  |  |  |  |
| Instrumental Activities of Daily Living Scale | 302<br>0.44<br>(1.55) | 177<br>0.38<br>(1.50) | 302<br>0.45<br>(1.35) | 177<br>0.42<br>(1.33) | 0.02<br>(-0.18 to -0.22) | 0.86 | 0.015038 |
| Mild Behaviour Impairment Scale | 302<br>7.31<br>(15.31) | 176<br>4.98<br>(13.61) | 303<br>6.92<br>(11.70) | 175<br>4.16<br>(7.11) | 0.52<br>(-1.71 to 2.75) | 0.65 | 0.073136 |
| EQ-5D-5L scale | 302<br>0.84<br>(0.16) | 174<br>0.84<br>(0.18) | 303<br>0.83<br>(0.16) | 175<br>0.85<br>(0.13) | 0.00<br>(-0.03 to 0.03) | 0.92 | 0 |
| EQ-5D-5L VAS | 302<br>76.1<br>(19.8) | 180<br>77.5<br>(18.1) | 303<br>76.4<br>(19.4) | 180<br>77.7<br>(18.1) | -0.7 (-3.7 to 2.4) | 0.69 | 0.04 |

### Impact of the START intervention on function, wellbeing and behaviour

In the non-cognitive secondary outcome measures, the treatment group also showed no significant benefit compared to the control group in function (P=0.86), behaviour (P=0.65) or wellbeing (P=0.92) (Table 3).

### Secondary analyses: Impact of supplementation in sub-groups of age, sex and vitamin D deficiency at baseline

Sub-group analyses showed no significant impact on trial outcomes in sub-groups of participants defined by age (five-year age brackets), gender or vitamin D deficiency severity, with the exception of differences between age groups on Paired Associate Learning (P = 0.02) (Table 4).

**Table 4:** Impact of vitamin D supplementation in sub-groups of age, sex and vitamin D deficiency severity (mild/moderate or severe)

| Subgroups | Age | Sex | Vitamin D deficiency |
| --- | --- | --- | --- |
|  | 50-59 vs. 60-69 vs.<br>70-79 vs. 80+ yrs | male vs.<br>female | Mild/moderate <50nmol/l vs.<br>severe <25 nmol |
| **Primary Outcome** |  |  |  |
| Trail making B | P=0.69 | P=0.79 | P=0.97 |
| **Secondary Cognitive Outcomes** |  |  |  |
| Verbal reasoning | P=0.13 | P=0.35 | P=0.95 |
| Paired Associate Learning | P=0.02 | P=0.48 | P=0.84 |
| Digit span | P=0.21 | P=0.83 | P=0.15 |
| Self-ordered search | P=0.13 | P=0.73 | Not estimable |
| Switching Stroop | P=0.42 | P=0.58 | Not estimable |
| **Secondary Non-Cognitive Outcomes** |  |  |  |
| IADL helplessness | P=0.21 | P=0.40 | Not estimable |
| IADL difficulty | P=0.52 | P=0.87 | Not estimable |
| MBI | P=0.08 | P=0.65 | P=0.65 |
| EQ-5D VAS | P=0.70 | P=0.14 | P=0.67 |
| EQ-5D utility score | P=0.37 | P=0.88 | P=0.92 |

### Per protocol analysis

Usable tablet count data were available in a subgroup of 256 participants. Adjusting for baseline versus follow-up tablet difference showed little or no impact on the treatment-control group effect of vitamin D at 24 months on the primary outcome (mean difference: -6174, 95% CI: -17095 to 4747, P=0.266).

### Compliance

49 participants were selected to complete a blood test analysis of vitamin D levels for compliance. Of these, 24 returned blood tests at both baseline and 24 months. All participants with blood level data in the intervention group showed improvement in vitamin D deficiency status at 24 months (P<0.001). Of the 256 participants with detailed tablet count data, the median compliance was 98%, and 98% of people had over 80% compliance with the intervention.

## Discussion

This study reports the findings from a remote randomised controlled trial of vitamin D supplementation in older adults with mild to moderate vitamin D deficiency and AACD. The results show that supplementation conferred no benefit to the primary outcome of executive function or to any secondary measures of cognition, function or well-being at the 24-month timepoint. The trial applied an appropriate double-blinded design and reached sufficient power to answer the research question, so this outcome is reliable and statistically significant.

There is already robust evidence for the benefits of supplementation in people with severe Vitamin D deficiency, but this study clearly shows the absence of benefit in individuals with mild to moderate deficiency, which is more prevalent in the population and less likely to be detected and treated than severe deficiency. The findings suggest that while there is a need to address low vitamin D intake for other health risks, such as osteoarthritis and osteoporosis, there may not be an impact on cognition as a result. There is increasingly clear evidence regarding strategies for promoting cognitive health in ageing, including physical activity, cognitive training, maintaining a healthy weight and managing key medical conditions such as depression, hypertension, diabetes and hearing loss. However, this trial indicates that vitamin D supplementation need not be included in this guidance for the public or clinicians when considering cognitive risk in individuals with moderate deficiency. The important exception is in individuals with severe vitamin D deficiency since this trial did not include sufficient individuals with a deficiency of <25nmol/l, as this was not the primary research question.

This study successfully delivered a fully remote randomised placebo-controlled clinical trial in 620 participants. It provides proof of concept for the innovative remote approach to trial delivery. Participants were recruited, treated, assessed and retained using fully remote methodology. Recruitment was achieved at the height of the Covid-19 pandemic in the UK when most clinical trials were halted and in a seven-month timescale that is unprecedented for traditional in-clinic trials. This was particularly successful due to the PROTECT-UK infrastructure, which allows targeted recruitment from the large ageing cohort with full consent for contact. The remote protocol for dispensing and tracking of study tablets was accurate and well-received by participants, and the study achieved good retention of participants across the 24-month period. Participants showed good compliance with the study protocol, as confirmed by the vitamin D blood level results, tablet count data and the completeness of the data. The online safety reporting approach enabled rapid, real-time collation and responsiveness to this low-risk intervention. Furthermore, the online and computerised assessment platform provided by the PROTECT-UK infrastructure facilitated high-quality data capture and depth of cognitive data without the need for in-person assessments. This study, therefore, provides important proof of concept for the remote trial design approach for low-risk interventions such as dietary supplements, nutraceutical products or repositioned drugs that do not require in-person physiological monitoring.

The study did, however, have some limitations. Recruitment was achieved through the online PROTECT-UK cohort, resulting in a self-selection bias towards females of Caucasian ethnicity with high educational attainment. The online nature of the trial also meant that participants were already fully digitally engaged. These design factors meant that the cohort was not fully representative of the UK population so caution should be taken in translating the findings to a population level, particularly considering the association of vitamin D and differing melatonin levels in different ethnic minority groups. This study was run for 24 months and showed no benefit to cognition so it is possible that a longer duration trial may have shown a more favourable outcome, although this would raise issues regarding the implementability of a long-term supplementation programme for public health.

This study provides robust evidence that Vitamin D supplementation does not result in cognitive benefits in individuals with mild to moderate deficiency. The findings indicate that while supplementation may be of value for other clinical indications and in people with severe deficiency, guidance does not need to include vitamin D supplementation for this patient group. Furthermore, the study provides robust proof of concept for remote clinical trial delivery using recruitment and engagement through the PROTECT-UK cohort platform and integrated computerised cognitive testing. This methodology should be considered for low-risk clinical trials.

## Data Availability

All data produced in the present study are available upon reasonable request to the authors

## Acknowledgements

This work was funded by the Jon Moulton Charitable Foundation. This paper represents independent research part-funded by the National Institute for Health and Care Research Exeter Biomedical Research Centre. The views expressed are those of the authors and not necessarily those of the NIHR or the Department of Health and Social Care. It was also supported by the NIHR Collaboration for Leadership in Applied Health Research and Care South-West Peninsula. JR receives funding from Alzheimer’s Research UK. The funders had no role in study design, data collection and analysis, decision to publish, or preparation of the manuscript. No funding has been provided by a pharmaceutical company or other agency to write this article. Authors have not been precluded from accessing data, and all authors accept responsibility for publication. https://www.exeter.ac.uk/

